# Automated Antibiogram Extraction from Unstructured Microbiology Reports: A Comparative Performance and Efficiency Analysis of Domain-Specific Named Entity Recognition (NER) Pipelines

**DOI:** 10.1101/2025.03.10.25323640

**Authors:** Mohamed Kamal, Omneya Hassanain

## Abstract

Antibiograms are essential tools in antimicrobial stewardship programs (ASPs), guiding empirical antibiotic therapy and tracking antimicrobial resistance (AMR) trends. However, the manual compilation of antibiograms from unstructured microbiology reports is labor-intensive and prone to delays. Here, we present a comparative study of three Natural Language Processing (NLP) approaches for automating data extraction from free-text reports: a rule-based Named Entity Recognition (NER) system, a statistical NER model using the spaCy library, and a transformer-based question-answering (QA) model leveraging DistilBERT. We generated a synthetic dataset of 3,000 microbiology reports to evaluate these methods, focusing on extraction accuracy (precision, recall, F1-score) and computational efficiency. The rule-based NER achieved perfect accuracy (F1 = 1.00) with minimal computational resources, making it highly suitable for real-time deployment. The spaCy model, after domain-specific fine-tuning, demonstrated strong performance (F1 = 1.00), effectively handling linguistic variations. In contrast, the transformer QA model showed moderate accuracy, excelling at extracting organism names but underperforming in detecting contamination status due to contextual ambiguities (F1 = 0.68-0.8). Computational efficiency analysis revealed that the rule-based and spaCy NER models could process reports rapidly with limited resources, while the transformer QA model required substantial computational power, potentially limiting its clinical utility. Additionally, we developed a prototype Expert System using R Shiny employing the rule-based NER to integrate extracted data into a real-time antibiogram dashboard, demonstrating the feasibility of these approaches in practical settings. The STEWEX (Stewardship Expert System) prototype has the capabilities of real-time building of fully functional antibiogram from simulated unstructured reports and simulated antibiotic susceptibility results. In conclusion, our results suggest that while advanced NLP methods offer flexibility, rule-based NER systems provide unparalleled accuracy and efficiency for data extraction from unstructured reports in ASPs, which represents bottle neck in development of antibiogram. Future efforts will focus on validating these approaches using real-world clinical data with the ultimate goal of fully automating antibiogram generation to support data-driven antimicrobial stewardship.

## Introduction

Antibiograms, which compile cumulative antimicrobial susceptibility data, are fundamental tools in antimicrobial stewardship programs (ASPs). They guide empiric antibiotic therapy, inform local resistance trends, and support formulary decisions. A well-maintained facility-specific antibiogram enhances empirical therapy selection, minimizes broad-spectrum antibiotic overuse, and contributes to longitudinal AMR surveillance, facilitating the early detection of drug-resistant pathogens and supporting infection prevention strategies [1,2]. Constructing an antibiogram typically requires extracting relevant information, such as organism identities and susceptibility results, from microbiology laboratory reports, which are often unstructured free-text narratives. While WHONET, a Windows-only open-access laboratory information system (LIS) for antimicrobial resistance (AMR) data analysis, effectively manages structured datasets, it lacks real-time entity extraction and classification capabilities and cannot process unstructured data.

Consequently, these limitations render it insufficient for fully automated antibiogram generation [3,4]. The free-text nature of these reports hinders their direct use in surveillance and decision support systems[5], making manual antibiogram compilation labor-intensive and time-consuming [6]. In large healthcare institutions, compiling data from hundreds and thousands of culture reports can take hours to days, creating a significant rate-limiting step in antibiogram generation[7,8]. Natural Language Processing (NLP) techniques offer a promising solution by converting unstructured text into structured data. In particular, *named entity recognition* (NER) can be employed to identify and extract key entities (such as organism names or interpretation notes) from narrative microbiology reports[6]. Prior studies have demonstrated the feasibility of NLP pipelines for microbiology data. For example, Yim *et al*. developed a system combining rule-based and statistical NLP to structure free-text culture reports, achieving an entity extraction F1-score of 0.889 [5]. More recently, Eickelberg *et al*. introduced an open-source rule-based tool (“MicrobEx”) that reliably parses microbiology reports (F1 >0.95)[9]. These works highlights the need for automated data extraction in this domain and the potential of NER-driven approaches. NER has been widely studied in the biomedical domain, leading to tools like MetaMap and MedCAT for general clinical text [10,11], and specialized methods for genes, diseases, and chemicals [12–14]. However, no existing system is optimized for microbiology reports, which present unique challenges such as: pathogen and strain-specific entity recognition, antimicrobial resistance marker extraction and structured antibiogram data parsing from free text. Existing NER tools are not optimized for these tasks, and while MicroBERT provides some microbiology adaptation, it lacks the capability for direct antibiogram generation[15] NER methodologies fall into two major categories: rule-based systems and machine learning-based models. The former relies on predefined linguistic patterns and domain-specific heuristics, offering deterministic accuracy and real-time execution with minimal computational overhead [16] In contrast, deep learning-based approaches, particularly those leveraging transformer architectures such as BERT and BioBERT, excel in resolving linguistic ambiguities by analyzing contextual cues within large corpora [17]. While transformer-based models such as BioBERT have demonstrated state-of-the-art performance in biomedical NLP, their high computational cost limits real-time deployment in hospital settings. Studies indicate that processing long clinical texts with transformer models requires substantial GPU/CPU resources, making them unsuitable for settings that need to process many reports quickly, such as hospital microbiology departments. Moreover, their memory and processing time requirements increase very rapidly as the text gets longer, further complicating their deployment for real-time antibiogram generation in resource-limited settings[18]. In this study, we focus on automating the rate-limiting step of antibiogram construction: extracting structured microbiological data from unstructured reports. We evaluate three different approaches to this task: A rule-based NER using pattern matching and domain-specific dictionaries, a statistical NER model using the spaCy library’s convolutional neural network (CNN) architecture, and a transformer-based question-answering (QA) model using a fine-tuned DistilBERT transformer. We compare these approaches in terms of extraction accuracy (precision, recall, F1-score) and efficiency. Our objective is to determine which method best automates data extraction for antibiogram compilation, thereby streamlining antimicrobial stewardship workflows.

## Methods

### Simulated Dataset Creation

We created a synthetic dataset of 3,000 simulated microbiology culture reports to develop and test the extraction methods. Each report was formatted to resemble a typical hospital microbiology report narrative, including the following unstructured fields: Organism identification, Time-to-positivity (TTP) and

Colonizer / Contaminant indicator. Times to positivity were simulated as random values (ranging 8–72 hours) to mimic laboratory observations. Contamination comments were inserted in 30% of reports. Reports were phrased with deliberate variations in organism names to test the robustness of extraction approaches. All 3,000 reports were then divided into development and evaluation sets (70% / 30% split). The development set was used to craft rules and train models, while the evaluation set was used for final performance assessment. Figure 1 illustrates the workflow with bottleneck step *“Extract structured Data from Reports”*. For the construction of the antibiogram, we also simulated antibiotic susceptibility results.

**Figure (1).**
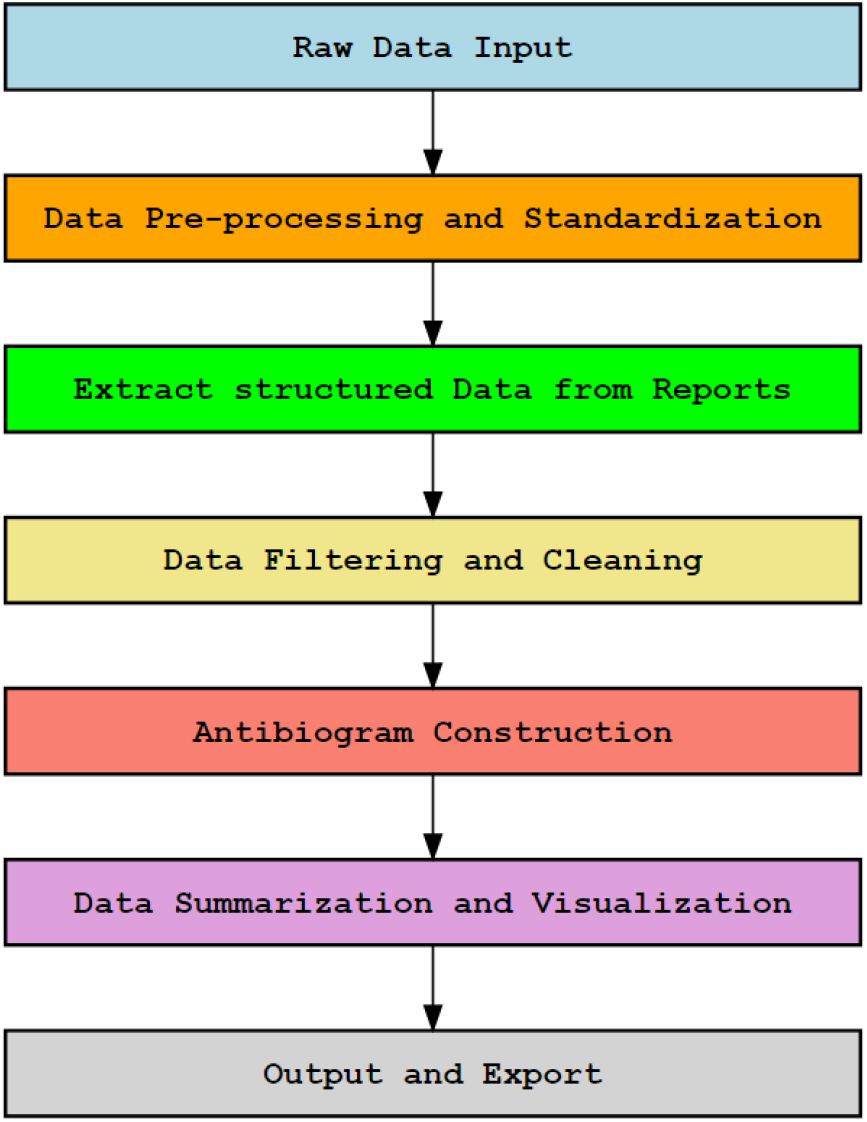
Expert System Workflow.

**Figure (2).**
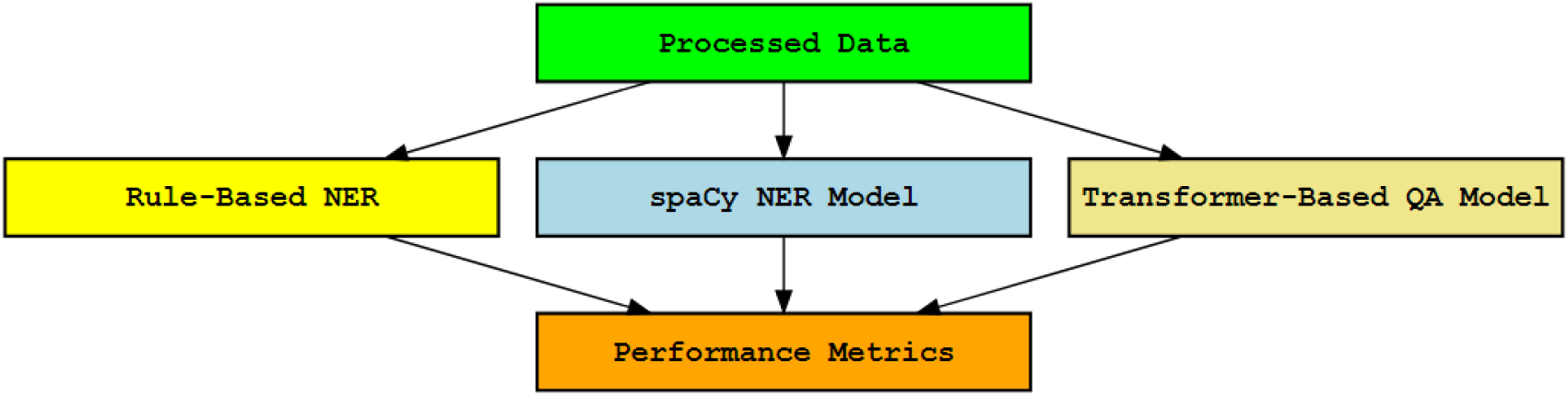
Approaches for Data Extraction.

In this workflow, raw data input initiates the pipeline by allowing the user to select and verify the integrity of their dataset via MD5 checksum comparison, ensuring authenticity before analysis. The system then proceeds with data pre-processing and standardization, removing duplicates and mapping columns to standardized nomenclature, including organism names and antimicrobial susceptibility test results. Next, structured data are extracted from the free-text fields and undergo a filtering and cleaning to exclude contaminants, colonizers, and redundant positive cultures, following the 14-day de-duplication rule recommended by the Clinical and Laboratory Standards Institute (CLSI) and used in CDC’s National Healthcare Safety Network (NHSN) protocols to prevent overestimation of resistance [19]. Once curated, the data supports the construction of an antibiogram, wherein patient collection locations are re-coded and susceptibility test values are categorized as sensitive, intermediate, or resistant. Subsequently, a data summarization and visualization step generates summary tables and heatmaps to convey the distribution of organisms and their antimicrobial susceptibility profiles. Finally, the workflow concludes with an output and export stage, producing reproducible files, including CSV summaries and PDF heatmaps, that document the entire process and facilitate further analysis or reporting.

### Approaches for Data Extraction

We implemented three approaches to extract the relevant entities (organism name, TTP, contaminant or colonizer) from the free-text reports:

#### 1. Rule-Based NER

We developed a custom rule-based parser leveraging regular expressions, string matching, and lookup dictionaries. This approach was built as follows: First, we compiled a dictionary of organism names (including common abbreviations) based on the NHSN Organism List, which incorporates SNOMED CT codes to standardize pathogen identification. This dictionary served as the foundational reference for extracting and categorizing organisms in microbiology reports [20].

The parser scans each report for any dictionary organism term (e.g. “Staphylococcus aureus” or “S. aureus”), using case-insensitive matching to account for capitalization. Next, a regular expression pattern identifies the TTP as a numeric value followed by “hour(s)” or “day(s)” for example the regex pattern *“(\\d+)\\s*(hour|hours|day|days)”*. This captures expressions like “14 hours” or “2 days” and converts days to hours for consistency. Finally, we detect contamination and colonization status by searching for keywords like “contaminant”, “contamination”, “colonizer” or phrases indicating no contamination. For example, the presence of “likely a contaminant” or “possible contaminant” is interpreted as a positive contaminant indicator, whereas “no contaminant” or absence of any such term is interpreted as a negative indicator. All rules were implemented in R using string operations and regular expression module. We iteratively refined the patterns on the development set to ensure they captured all variants present. The rule-based system does not require any training data beyond the predefined patterns and lists, and its output is entirely deterministic.

#### 2. spaCy NER Model

We employed spaCy (v3.2), an NLP library, to train a statistical NER model for our task. By default, spaCy’s NER uses a transition-based architecture with a convolutional neural network (CNN) that can learn to recognize custom entity types given annotated examples [21]. We defined four custom entity labels – ORGANISM, TTP, COLONIZER and CONTAMINANT – corresponding to the information to extract. For model initialization, we experimented with spaCy’s English core model pre-trained on general text, which provides baseline language features. We then fine-tuned this model on our domain-specific corpus. Using the development dataset, we manually annotated each report with the spans corresponding to the organism name, the TTP phrase, and the contaminant indicator phrase. For instance, in *“Blood culture grew Escherichia coli. Time to positivity 18 hours. Possible contaminant*.*”*, the spans “Escherichia coli” would be labeled ORGANISM, “18 hours” as TTP, and “Possible contaminant” as CONTAMINANT. A total of 2,100 reports were annotated in this manner for training, and spaCy’s training pipeline was run for 30 epochs. We used a dropout rate of 0.2 to prevent overfitting by randomly deactivating neurons during training, and the Adam optimizer due to its efficiency in handling sparse gradients and adaptive learning rates. Early stopping based on development-set performance was implemented to prevent overfitting by halting training when performance ceased to improve [22,23]. After training, the model was applied to the evaluation set to extract entities. We also evaluated spaCy’s out-of-the-box performance *before* fine-tuning (i.e. using the pre-trained model without additional training on our data) to serve as a baseline.

#### 3. Transformer-Based QA Model

For the third approach, we defined the extraction task as a question-answering problem using a transformer model. We fine-tuned DistilBERT (a distilled version of BERT chosen for its smaller size and faster inference),[24] on a set of question-answer pairs derived from our reports. We used question templates designed to extract the organism name, the time-to-positivity (TTP), and the status of the isolate in terms of contamination or colonization. These templates were intended to prompt the model to identify specific spans of text directly from the reports, such as the organism type, a time measurement, or terms indicating contamination or colonization. We fine-tuned DistilBERT on these QA pairs (6,300 pairs from 2,100 reports, since each report yields 3 questions) for 3 epochs using Hugging Face’s Transformers library in Python[25]. The model learned to predict answer spans within the report text based on the context provided by the report text itself, guided by the prompts from the question templates.

We also evaluated the performance of the pre-trained QA model (DistilBERT fine-tuned on SQuAD for general QA) on our evaluation set questions without additional training. This comparison was made to assess the benefits of domain-specific fine-tuning for handling specialized information effectively.

##### Evaluation Metrics

We evaluated each approach’s extraction performance using standard NER metrics:

- **Precision:** the proportion of entities extracted by the system that were correct (relevant to the report).
- **Recall:** the proportion of relevant entities in the report that were successfully extracted by the system.
- **F1-score:** the harmonic mean of precision and recall, providing a single measure of accuracy.

These metrics were computed for each entity type (organism, TTP, contaminant and colonizer) per approach. An extraction was considered *correct* if the text span exactly matched the gold standard answer for that field (with minor variations in numeric format allowed for TTP, e.g. “1 day” vs “24 hours” counted as a match). For the contaminant / colonizer indicator, any correctly identifying the presence or absence of contamination / colonization was counted as correct. We calculated precision, recall, and F1 both before and after fine-tuning for the ML approaches (spaCy NER and transformer QA) to quantify improvement. The rule-based method does not involve a learning phase, but we treated its initial development iteration as a “before” state and its final optimized form as “after” to allow comparison. In addition to accuracy, we also assessed computational efficiency.

We measured the processing time required by each approach to parse the entire evaluation set. All experiments were run on cloud computing instances in a virtualized Linux environment (kernel version 6.10.2-x86_64-linode165) on a KVM-based platform. The underlying hardware featured an AMD EPYC 7542 32-Core Processor (32 CPUs) and 64 GiB of RAM. No dedicated GPU was present, thus, all model inferences, including those for the transformer model, were executed on the CPU. We also monitored CPU and memory usage, as well as practical issues such as model loading time, because these factors critically impact deployment in clinical settings.

## Results

### Extraction Accuracy

After development and fine-tuning, the rule-based system and both machine learning models were applied to the 900 unseen evaluation reports. Table 1 summarizes the precision, recall, and F1-score achieved by each method, both before any domain-specific tuning (initial) and after fine-tuning or rule optimization (final).

**Table 1.** Presents the precision, recall, and F1-scores (broken down by entity type) for each extraction method before and after their respective training or rule-building steps. As shown, spaCy exhibits 0% performance across all categories prior to domain-specific training, but after fine-tuning, it achieves 100% F1 for organisms, colonizer, contaminant, and TTP. The QA approach initially shows perfect F1-scores (100%) for organism and TTP extraction but struggles with colonizer (59%) and contaminant (23%). After fine-tuning, QA improves considerably in colonizer (68%) and contaminant (80%) detection while retaining 100% F1 for organism and TTP. The rule-based NER similarly starts at 0% for all entities before the pattern dictionary is built, but reaches 100% across all metrics once the dictionary and rule optimization is finalized.

| Method | Organism<br>P | Organism<br>R | Organism<br>F1 | Colonizer<br>F1 | Contam.<br>F1 | TTP<br>F1 |
| --- | --- | --- | --- | --- | --- | --- |
| **spaCy Before Training** | 0% | 0% | 0% | 0% | 0% | 0% |
| **spaCy After Training** | 100% | 100% | 100% | 100% | 100% | 100% |
| **QA Before finetuning** | 100% | 100% | 100% | 59% | 23% | 100% |
| **QA After finetuning** | 100% | 100% | 100% | 68% | 80% | 100% |
| **Rule Based NER before PDB / RO** | 0% | 0% | 0% | 0% | 0% | 0% |
| **Rule Based NER after PDB / RO** | 100% | 100% | 100% | 100% | 100% | 100% |

\*PDB RO: pattern dictionary building / rule optimization

To gauge how these methods might perform on real-world data, we compare our results to those reported in the literature. Yim *et al*. (2015) reported an entity extraction F1 of 0.889 on actual hospital culture reports using a hybrid NLP system [5]. Our learned models (spaCy and QA) exceeded this on simulation, but real-world noise could reduce their performance. Meanwhile, a recent rule-based system (MicrobEx) achieved F1 >0.95 in multi-center validation [9], which is comparable to our rule-based outcome.

### Computational Efficiency

The three approaches demonstrated marked differences in runtime and resource usage on our cloud instance (AMD EPYC 7542, 32 cores, 64 GiB RAM) running a virtualized Linux environment.

The rule-based NER system was exceptionally lightweight. It processed all 900 test reports in under 0.5 seconds on a single CPU thread - roughly 1800 reports per second - with a small memory footprint. This speed, driven by simple string-matching and regular expression operations, easily scales to thousands of reports in real time on standard hardware.

The spaCy NER model also performed efficiently. On CPU, it processed the 900 reports in about 1.9 seconds (approximately 470 reports per second). The initial model loading took around 2 seconds—a one-time cost—after which the optimized CNN architecture enabled rapid entity recognition. Memory usage remained moderate due to the loaded model and word embeddings. For this scale, CPU execution provided sufficient throughput, ensuring that daily report volumes in a large hospital could be handled with minimal latency.

In contrast, the transformer-based QA approach was the most resource intensive. On CPU, DistilBERT took roughly 28 seconds to answer three questions per report across the 900 reports, yielding an effective throughput of about 32 reports per second, or roughly 96 individual QA inferences per second. This slower performance is attributable to the higher computational demands of transformer models and the need for multiple passes over each report (one pass for organism extraction, another for TTP, and a third for contamination / colonization status). Additionally, the QA model’s memory footprint was substantial. Although transformer models typically benefit from GPU acceleration, our experiments were conducted on CPU, and under these conditions the transformer was approximately 10× slower than the spaCy model. While processing 32 reports per second may be workable for many labs in a batch setting, the increased latency and computational cost could be challenging for real-time or resource-limited deployments.

One compelling reason to continue using a rule-based NER system is the significant overhead associated with training a spaCy model. Although spaCy achieves excellent performance after fine-tuning, the process of annotating training data and optimizing the model is both time- and resource-intensive. In contrast, the rule-based approach requires no training phase, enabling immediate deployment and simpler maintenance. We also developed tools for facilitated dictionary building and enhanced rules optimization using R. Furthermore, its transparent logic allows for quick adaptations to changes in report formats or terminology without the need for re-training, making it particularly attractive in dynamic clinical environments where rapid updates are essential.

### Usability and Integration

Beyond raw accuracy and speed, we evaluated how *usable* and easily integrable each solution would be in a clinical setting. The rule-based system produces very predictable outputs. The results are easily interpretable by clinicians and microbiologists. Any errors can be traced to specific rules and corrected by updating the pattern or dictionary. This transparency is advantageous for clinical adoption, as stakeholders often prefer to understand *why* a system produces a certain output. Integrating the rule-based extractor into the lab workflow would be straightforward. Given its small footprint, it can run on existing LIS (Laboratory Information System) infrastructure without minimal preparation and hardware.

The spaCy NER model offers a balance of good performance with some loss of transparency. Once trained, it acts as a black box that labels text spans without explaining its reasoning. In practice, this means validation and debugging are harder as the operator cannot easily pinpoint *why* the model missed an entity except by examining the training data. However, the spaCy model’s errors in our study were infrequent and sometimes obvious (e.g. it might output an entity spanning an extra word). Users can correct outputs manually if needed or retrain the model with additional examples. From an integration standpoint, deploying spaCy would require installing the Python environment and model on a hospital server. One advantage of the ML approach is that it may generalize better to slight variations in language than a rigid rule-based system. For instance, if new wording or new organism names appear, a trained model might handle them if they resemble known patterns.

The transformer QA approach is the most complex to integrate. It involves orchestrating multiple question queries per report and then processing the answers. This complexity could introduce failure points (e.g., if one query fails or returns an unexpected string). In our prototype, we had to implement post-processing rules for the QA outputs, such as mapping an answer like “no” or “none” in response to the contaminant question to a standardized “No contamination” label. Essentially, the QA approach may still require a layer of rules to interpret its answers in a structured format. Users might find the QA outputs slightly less straightforward, since they come as free text answers. For instance, for a given report, the model might output: Q1 -> “Streptococcus pneumoniae”, Q2 -> “36 hours”, Q3 -> “contaminant”. These are correct, but the third answer might sometimes be just “possible” or “likely”, needing translation into a consistent flag. Despite these challenges, a QA system has a unique flexibility: one could ask additional questions without retraining the model (e.g. “What sample type is this culture from?”) as long as the model has learned to find that kind of information. This could make the system extensible. In our integration tests with the Shiny app, the QA model was the only one that had noticeable latency (a few seconds delay when analyzing a batch of reports), whereas the rule-based and spaCy models felt instantaneous to the end user.

In terms of clinical workflow, and apart from the differences in performance metrics, all three approaches ultimately provide the same desired output: structured data (organism, TTP, contaminant / colonizer flag) that can be fed into an antibiogram development pipeline. We simulated this by having each method populate a database table with columns for organism and an indicator if it was a true pathogen (non-colonizer / non-contaminant). The antibiogram construction then simply counts each organism that is a true pathogen to compute prevalence and susceptibility (if susceptibility data were included, that would be aggregated per organism). We found that when the extraction was accurate, the downstream antibiogram matched the ground truth. When extraction errors occurred (mostly in the QA model’s outputs), the antibiogram could be slightly off e.g. an organism missed by the extractor would be under-counted.

### Discussion Key Findings

In this study, we compared three NLP-based approaches to automate the extraction of structured data from unstructured microbiology reports as a step toward automated antibiogram generation. Our results demonstrate that, with sufficient domain-specific training, a statistical NER model like spaCy can match the accuracy of expert-defined rules. The spaCy model, after extensive fine-tuning, achieved an F1-score of 1, indicating that machine learning models can effectively recognize clinical entities given sufficient domain-specific training data. This improvement highlights the importance of adapting NLP models to the specialized language used in microbiology reports.

The rule-based NER approach provided unparalleled accuracy with minimal computational cost, achieving 100% precision and recall on the test data. This suggests that, in settings with consistent report formats, a carefully crafted rule-based system can effectively eliminate extraction errors and is on par with state-of-the-art tools in terms of accuracy, at least for the targeted entities. This aligns with recent findings by Eickelberg *et al*., who showed a rule-based algorithm could attain F1 >0.95 for culture report parsing across multiple hospitals [9].

The transformer QA model presented a mixed outcome. It excelled at extracting well-defined entities, particularly organism names, but struggled with other less direct classification tasks, such as determining contamination status. Its lower F1-score of 0.68-0.8, particularly for contamination / colonization detection, highlights that transformers, despite their advanced contextual understanding, may require additional rule-based heuristics or further optimization to handle certain tasks reliably. The transformer QA’s uneven performance is a caution that the latest NLP architectures are not guaranteed to outperform older methods, particularly if the task (e.g. yes/no classification in context) is somewhat outside the typical scope of the model’s training. Overall, all three approaches succeeded in automating data extraction to a large degree, with the rule-based and spaCy NER being the most reliable in our evaluation.

### Clinical and Computational Considerations

Choosing the right approach for real-world deployment involves balancing clinical requirements and computational resources. Rule-based NER offers unparalleled accuracy with minimal computational cost, making it ideal for hospitals with limited IT infrastructure. Its transparency is a key advantage, allowing clinicians to understand and modify rules easily as clinical guidelines evolve. However, its reliance on fixed patterns makes it less flexible; report format changes or unexpected entities can cause failures if rules were not re-visited. Maintenance is straightforward but requires expert input to update rules when necessary. Given that hospital microbiology reports and organism nomenclature evolve slowly, a rule-based system can remain accurate over long periods with minimal intervention.

The spaCy NER approach requires an initial investment in annotating training data, which can be labor-intensive. However, a well-annotated corpus can be reused or shared across institutions, making the effort more worthwhile. Unlike rule-based systems, spaCy can handle linguistic variations more robustly; for example, it can learn different phrasings such as “contamination likely” and “likely contaminant” without needing explicit rules for each. In our results, spaCy occasionally mis-labeled entities but managed to generalize well to novel phrasings, demonstrating effective learning from training examples.

From a computational perspective, spaCy is optimized for efficiency and can run on standard hardware. The requirement for a Python environment is a minor challenge but can be addressed with containers or standalone executables approved by hospital IT departments. However, maintaining accuracy requires periodic retraining if report formats or lab procedures change, unlike the straightforward rule adjustments needed for rule-based systems. This highlights a trade-off: rule-based NER demands less frequent but expert-led updates, while spaCy requires ongoing retraining to remain accurate.

The transformer QA model offers deep contextual understanding and flexibility, allowing different information to be extracted by simply changing the question without retraining. For instance, it can extract a quantitative measure by responding to questions like “How many colony-forming units were reported?” This adaptability is valuable in expert systems for extracting diverse insights from text. However, in practice, the QA model required extensive fine-tuning and did not consistently outperform simpler methods for core entities. Its computational cost is a significant drawback, often requiring a GPU or accepting slower performance, which can be a barrier for some ASP teams.

A notable concern is the model’s tendency to produce overconfident but potentially incorrect answers, which might mislead non-experts. For example, if it incorrectly labels an organism as a “contaminant” instead of a true pathogen, clinicians might mistakenly exclude it from the antibiogram. This risk emphasizes the need for reliable systems or a human-in-the-loop for verification. Moreover, the QA model would likely require a validation step for each output to ensure accuracy, reducing its practical advantage. This trade-off between flexibility and the need for careful oversight highlights the importance of context-specific deployment decisions.

From a clinical impact perspective, even partial automation can significantly speed up antibiogram preparation. By automatically extracting basic microbiology results, the stewardship team can focus more on analysis and interpretation rather than manual data processing. Our findings suggest that a rule-based or well-trained ML system could be integrated into the lab results pipeline to enable continuously updating antibiograms. This could allow for more frequent updates (e.g., quarterly or monthly instead of annually) by eliminating data aggregation as a bottleneck. Timelier antibiograms can support more responsive antimicrobial prescribing practices by providing clinicians with the latest resistance patterns [9,27] Additionally, structuring organism and susceptibility data is a critical first step toward advanced analytics. Once in a structured format, these data could be leveraged for statistical analyses or machine learning to detect trends, outliers, or outbreaks in near real-time [9].

### Limitations

Our study has several limitations. First, it relied on simulated data rather than real hospital reports, which may not fully capture the variability and noise of actual clinical documentation, such as typos, rare organism names, and multi-organism infections. To ensure accuracy before deployment, our systems will need to be validated using real reports from our clinical microbiology lab. We anticipate that the rule-based system might require additional rules for edge cases, while the spaCy and QA models may experience a slight drop in accuracy with unseen phrasing. However, similar studies give us confidence; for instance, a hybrid NLP system achieved around 84% sensitivity in extracting antibiotic susceptibilities and contamination status from hospital reports [2] Additionally, our rule-based approach is expected to perform similarly to MicrobEx, which has been validated externally [9].

Second, our evaluation was limited to a specific set of entities; organisms, TTP, and contamination / colonization status, essential for antibiogram statistics. However, a complete solution must also extract antibiotic susceptibility results. This task is more complex as it involves identifying drug names and interpreting results (S/I/R or MIC values). Previous systems have shown promise in this area; for example, Matheny et al. (2009) reported a 96% positive predictive value in mapping antibiotics to bacteria using a rule-based approach [2] In our prototype described below, we extended our system to extract these results from simulated data. Still, the limitation of using simulated data applies here.

Lastly, our study did not cover long-term maintenance and evaluation. To ensure sustained accuracy, we propose periodic spot-checks—randomly sampling 5–10% of extracted antibiogram entries each month for verification against original reports. Additionally, incorporating user feedback via a Shiny dashboard could allow clinicians to flag errors for review, helping to catch issues early.

## Conclusion

We developed and evaluated an expert system for automated extraction of microbiology data to build cumulative antibiograms, comparing rule-based NER, spaCy NER, and a transformer-based QA model. Our findings show that a rule-based approach, despite its simplicity, provided a scalable and highly accurate solution for this task, achieving 100% accuracy on simulated reports with minimal computational demands. The spaCy machine learning model, after fine-tuning on domain-specific data, also performed very well (100% F1), demonstrating that NLP models can be adapted to recognize clinical microbiology entities with high precision. The transformer QA model, while powerful in theory, was less consistent in our application, it excelled at direct entity extraction (organism names) but struggled with interpreting contamination and colonization status. Automating the construction of antibiograms can have a meaningful impact on antimicrobial stewardship. By eliminating the manual data processing bottleneck, hospitals can generate up-to-date antibiograms more efficiently and frequently, enabling clinicians, pharmacists, ID teams and supply chain staff to make informed antibiotic choices based on the latest local resistance data. Our proposed rule-based system in particular could be integrated into laboratory information systems to provide real-time updates to an antibiogram dashboard whenever a new culture result is finalized. This kind of expert system integration aligns with the goals of ASPs to use data-driven decision support to improve antibiotic use [27]. Moreover, structured data outputs can feed into surveillance systems and alert programs - for instance, automatically flagging when a normally rare pathogen suddenly appears more frequently.

### STEWEX (The Antimicrobial Stewardship Expert System)

We developed a prototype Expert System in R Shiny, implementing a rule-based NER approach to integrate extracted data into a real-time antibiogram dashboard. This STEWEX prototype demonstrates the feasibility of building fully functional antibiograms in real time using simulated unstructured reports and simulated antibiotic susceptibility results. Figures (3) and (4) illustrate how STEWEX processes data and presents real-time antibiogram insights, highlighting the practical benefits of a rule-based NER engine.

**Figure (3)(a).**
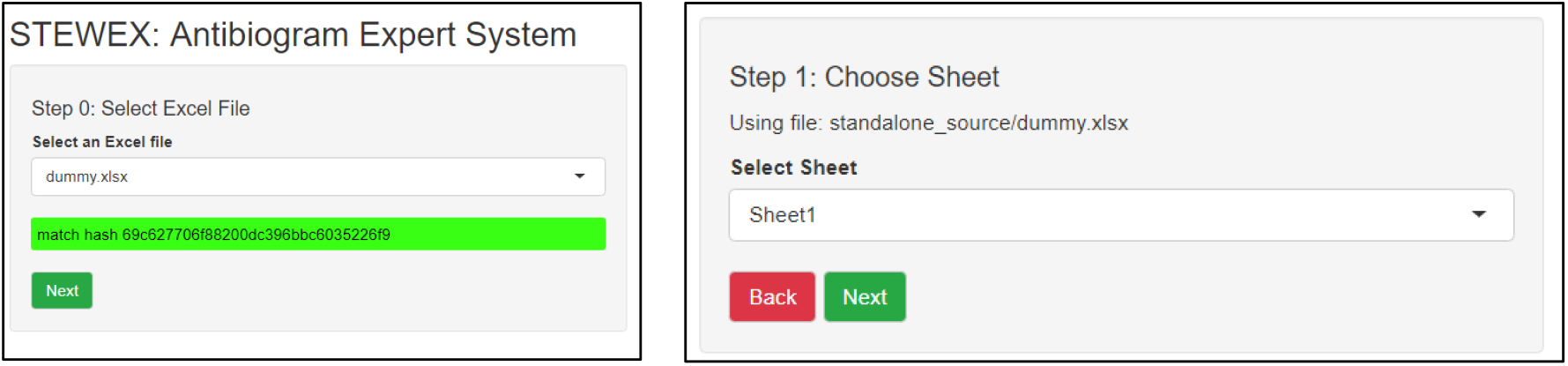
STEWEX data selection interface.

The user-friendly STEWEX interface starts by letting the user choose from available source files. In real-world scenarios where integration with the Hospital Information System takes place, the files containing the raw data for the antibiogram would undergo a checksum validation and be integrated with the Expert System. Here, the stored hash matched selected file ensuring file integrity. To minimize efforts on users especially the health informatics team, STEWEX has the capability to map the columns of the integrated report to the required columns to build the antibiogram, eliminating the need to rename columns and make the system malleable to any available data input.

Moreover, STEWEX also allows for defining data types of key fields required to construct the antibiogram. See Figures (3) (b) and (c) for more details. The final step is to run the analysis and export output files. By default, STEWEX exports seven files, including a processed data sheet, an operation log, an organism distribution by location, and a susceptibility heatmap. Additional reports can be customized as needed. The system also outputs operational aids and data quality indicators (e.g., MRN pattern checks for different patient categories and culture ID validation).

**Figure (3) (b).**
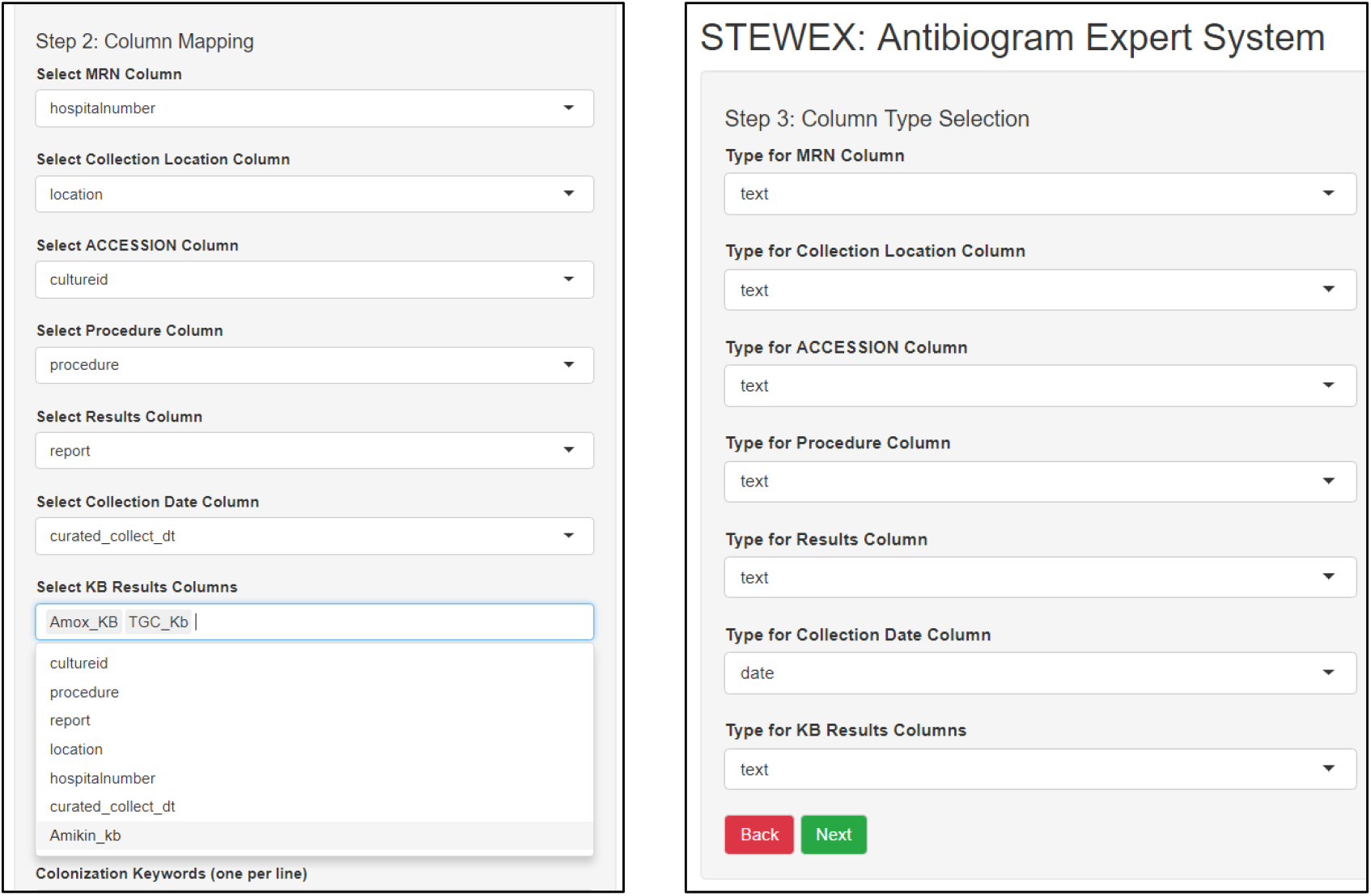
Mapping the required fields to the dataset fields (c) Defining data types.

**Figure (3) (d).**
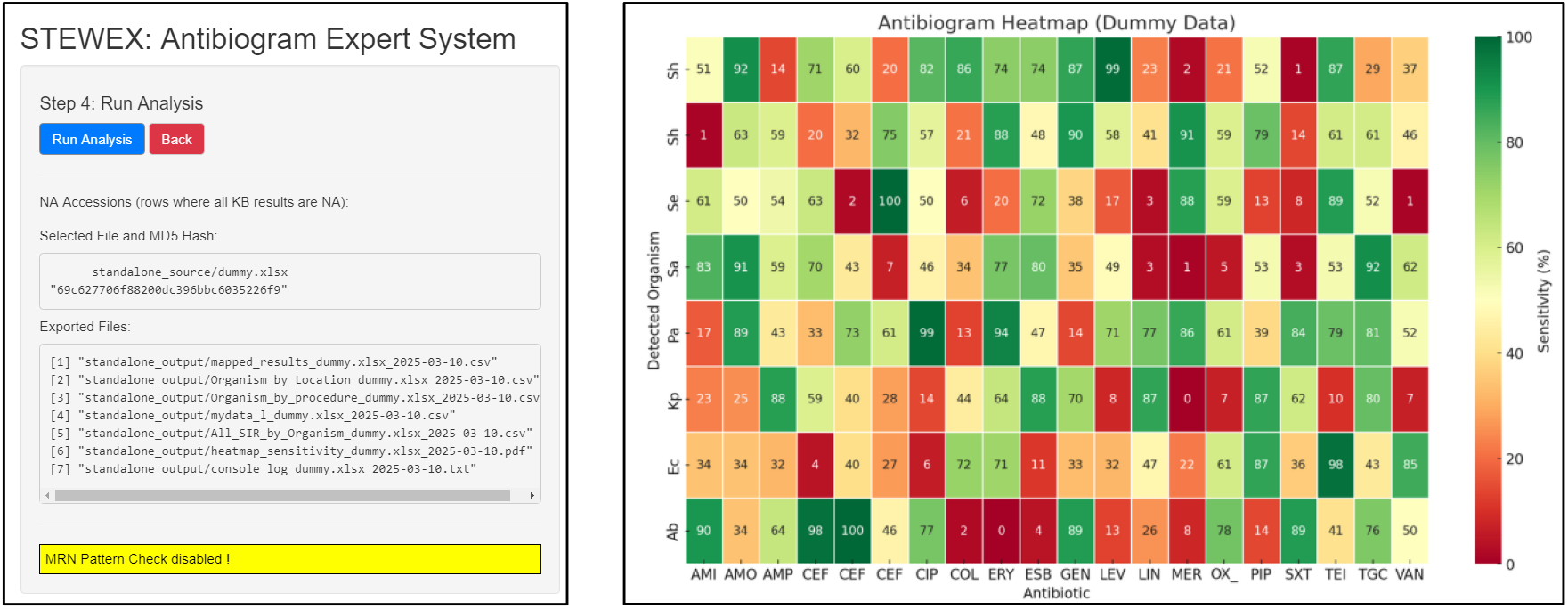
Running Analysis and exporting output (e) Antibiotic susceptibility heatmap.

Figure (4) illustrates a key background operation of the rule-based NER engine, showing how STEWEX identifies organisms, colonization status, contamination, time to positivity, and multiple detections, ultimately organizing these findings into structured data frames.

**Figure (4).**
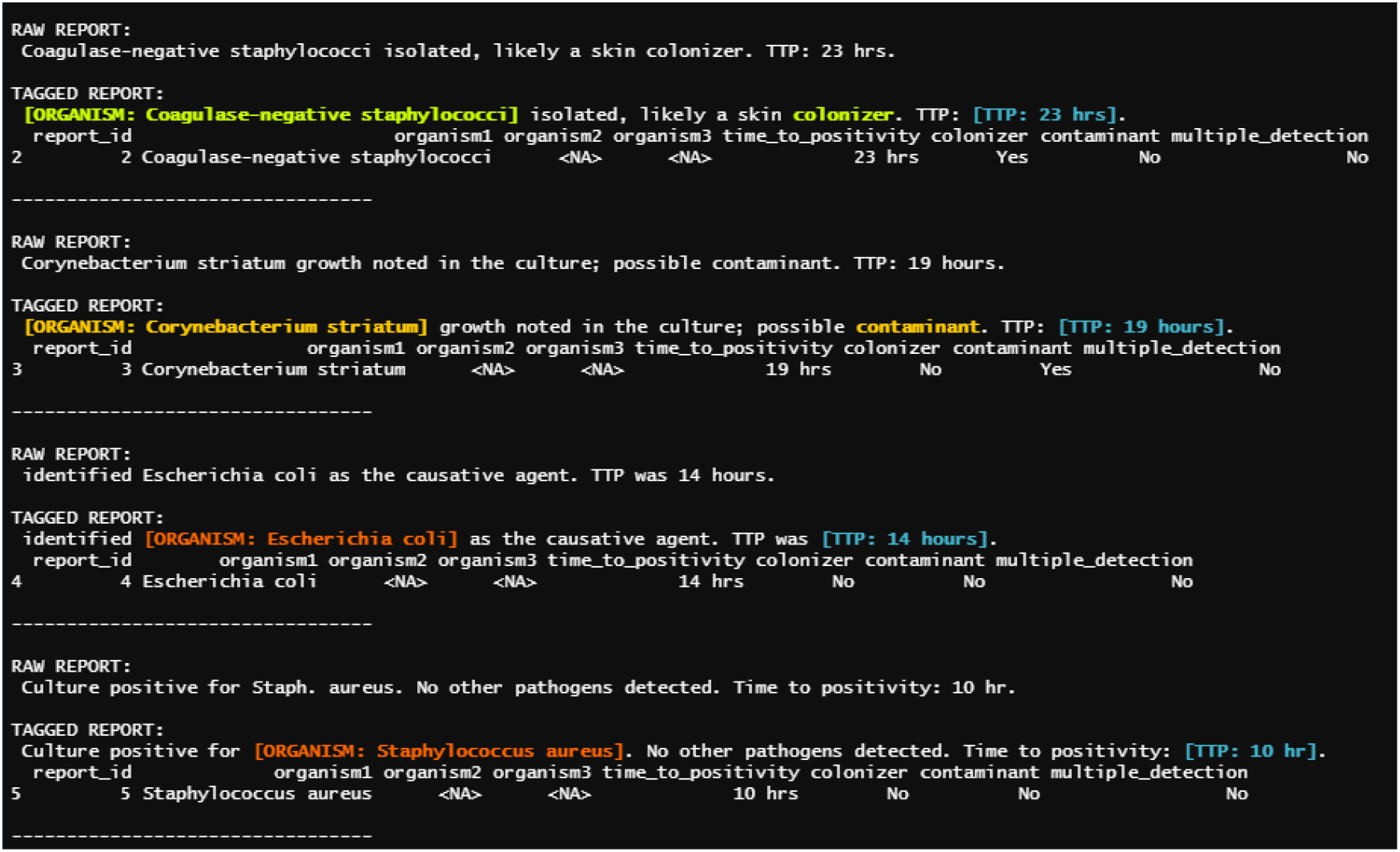
Processing of Raw Reports.

In conclusion, this study supports the feasibility of an automated, NLP-driven approach to antibiogram creation. Each technique has its pros and cons: rule-based NER offers unmatched precision and speed in well-defined settings, statistical NER (spaCy) provides adaptability with high accuracy given the right training, and transformer QA offers flexibility but may require further optimization for niche tasks. Because transformer models are designed to handle highly ambiguous language, and medical documentation tends to be more straightforward, their advantages do not compensate for the slower speed compared to rule-based NER. Ultimately, our work contributes to the larger vision of leveraging artificial intelligence and expert systems in healthcare to streamline processes and support the fight against antimicrobial resistance.

## Data Availability

All data used in the study were simulated using R version 4.2. The simulation script is available upon reasonable request to the authors

